# Community end-of-life care during the COVID-19 pandemic: Initial findings of a UK primary care survey

**DOI:** 10.1101/2021.02.15.21251756

**Authors:** Sarah Mitchell, Phillip Oliver, Clare Gardiner, Helen Chapman, Dena Khan, Kirsty Boyd, Jeremy Dale, Stephen Barclay, Catriona Mayland

## Abstract

**Background:** Thousands of people in the UK have required end-of-life care in the community during the COVID-19 pandemic. Primary healthcare teams (general practice and community nursing services) have provided the majority of this care, alongside specialist colleagues. There is a need to learn from this experience in order to inform future service delivery and planning.

**Aim:** To understand the views of general practitioners and community nurses providing end-of-life care during the first wave of the COVID-19 pandemic.

**Design and Setting:** A web-based, UK-wide questionnaire survey circulated via professional general practice and community nursing networks during September and October 2020.

**Method:** Responses were analysed using descriptive statistics and an inductive thematic analysis.

**Results:** Valid responses were received from 559 individuals (387 community nurses, 156 General Practitioners (GPs) and 16 unspecified role), from all regions of the UK. The majority reported increased involvement in providing community end-of-life care. Contrasting and potentially conflicting roles emerged between GPs and community nurses. There was increased use of remote consultations, particularly by GPs. Community nurses took greater responsibility in most aspects of end-of-life care practice, particularly face-to-face care, but reported feeling isolated. For some GPs and community nurses, there has been considerable emotional distress.

**Conclusion:** Primary healthcare services are playing a critical role in meeting increased need for end-of-life care in the community during the COVID-19 pandemic. They have adapted rapidly, but the significant emotional impact, especially for community nurses, needs addressing alongside rebuilding trusting and supportive team dynamics.

**How this fits in (4 sentences):**

- This study provides insights into experiences of delivering end-of-life care in the community during the first wave of the COVID-19 pandemic from the perspectives of UK general practitioners (GPs) and community nurses.
- Services have changed and adapted rapidly to meet increased need in terms of both volume and complexity, with community nurses taking greater responsibility for most areas of palliative care clinical practice, and GPs undertaking more care planning conversations.
- While GPs and specialist palliative care services conducted more remote consultations, community nurses carried out face-to-face end-of-life care and reported a feeling of isolation.
- As the pandemic progresses, and the increased need for end-of-life care in the community persists, more effective service models and multi-disciplinary teamwork in primary care are urgently needed.

## Introduction

The COVID-19 pandemic has caused primary healthcare services (general practice, and community nursing services including district nurses) to dramatically change their traditional models of service delivery over a short timeframe. Palliative and end-of-life care for people at home was affected from the outset (1-3). During the first 10 weeks of the pandemic there was a significant increase in deaths in the community, particularly in people aged over the age of 75 years. Deaths in care homes increased by 220% and deaths at home increased by 77%, while deaths in a hospice fell by 20% (4), presenting significant workload in this area for primary healthcare teams. While guidance documents for both general practice and community nurses outlined end-of-life care as urgent priorities (5, 6), there were many new challenges in the delivery of that care, including the need for more remote consultations and the use of personal protective equipment. Furthermore, there were concerns about drug and equipment supplies and the need to manage new symptom profiles associated with COVID-19 (7). Previous research and policy guidance for primary healthcare services in pandemics do not tend to refer to community end-of-life care, and there was little evidence to inform and guide the necessary service changes (8, 9).

There is therefore an urgent need to understand more about the role and response of general practice and community nursing services in the delivery of palliative care during the COVID-19 pandemic in order to inform practice and policy through future phases of COVID-19 and similar pandemics.

### Aims

The study aimed to provide insights and understanding into the experiences and perceptions of primary healthcare professionals within the United Kingdom (UK) providing palliative and end-of-life care in the community during the first wave of COVID-19, including how services changed and adapted.

## Method

An online questionnaire study was the most feasible method to meet the research aims, gathering both quantitative and qualitative data rapidly. The STROBE checklist informed the study design (10). An online survey instrument was devised, informed by patient and public involvement, a literature review conducted by this research team (9), the CovPall study of palliative care services (11) and feedback from a study advisory group of clinicians and commissioners. The survey instrument was pre-tested by a group of 17 general practitioners (GPs) and community nurses.

The survey contained open and closed questions, to collect demographic data about the participants and the services they work in, then qualitative and quantitative data about specific aspects of community palliative and end-of-life care provision during COVID-19, including changes in services, challenges and exemplars of good practice.

### Patient and public involvement

Patient and public involvement (PPI) was a key component of this research, with a PPI co-applicant, joining the research team, and further PPI sought through the University of Sheffield Palliative Care Studies Advisory Group (PCSAG). Both our local PPI work and a national consultation exercise (12) highlighted the importance of the provision of end-of-life care in the community during COVID-19. Group members provided comments on the literature review and assisted in the development of the research questions, survey instrument and overall design of this research.

### Data collection

The survey was circulated via social media and UK professional networks locally and nationally, including the Royal College of General Practitioners, the Society for Academic Primary Care, the Royal College of Nursing, the Queen’s Nursing Institute and the National District Nursing Network and via social media. Responses from GPs or community nurses were included, responses from other healthcare professionals were excluded. The aim was for a sample size of 500, having previously achieved sample sizes in this region (13). Data was collected between 1^st^ September and 16^th^ October 2020. All responses were anonymous.

### Data analysis

Quantitative data were analysed using descriptive statistics using SPSS (version 26). Qualitative responses were anonymised, uploaded into NVivo 12 software (version 10), and analysed using an inductive, iterative thematic approach (14). For the purposes of this paper, the qualitative findings relating to personal reflections about the provision of end-of-life care during the initial wave of the COVID-19 pandemic are reported. Codes were assigned to each data item, then all codes collated, mapped out, and compared to extrapolate overarching themes. The data analysis was led by SM, CR and PO with regular discussion of the emerging findings in order to reduce lone researcher bias (15).

## Results

### Demographics

In total, 563 responses were received; healthcare professionals out with our target group were excluded, leaving 559 valid responses. Of these, there were 387 community nurses, 156 GPs and 16 who did not specify their role. The latter group was included in summary descriptive statistics but omitted from any group comparisons or qualitative analysis. (Table 1). Respondents represented all countries within the UK with over three-quarters (77.1%) working in England. Different types of rurality were represented with the largest proportion from a ‘mixed urban and rural’ area (222, 39.9%). Results are presented in this section as three inter-related themes.

**Table 1:** Demographic details of respondents and details about provision of care for dying patients during the pandemic (n=559)

|  | N | % |
| --- | --- | --- |
| **Which country do you work in? (n=559)** |  |  |
| England | 431 | 77.1 |
| Scotland | 65 | 11.6 |
| Wales | 47 | 8.4 |
| Northern Ireland | 16 | 2.9 |
| *Missing* | - |  |
| **What type of area do you work in mainly? (n=556)** |  |  |
| Mixed urban and rural | 222 | 39.9 |
| Urban | 179 | 32.3 |
| Rural | 106 | 19.0 |
| Inner city | 49 | 8.8 |
| *Missing* | 3 |  |
| **What is your role? (n=543)** |  |  |
| **Doctor** | **156** | **28.7** |
| GP Partner | 104 | 19.1 |
| Sessional GP | 45 | 8.3 |
| Other e.g. GP in training | 7 | 1.3 |
| **Community nurse** | **387** | **71.3** |
| Community Staff Nurse | 150 | 27.6 |
| District Nurse (including team leaders) | 159 | 29.3 |
| Advanced Nurse Practitioner | 32 | 5.9 |
| Community Matron | 24 | 4.4 |
| Community Healthcare Assistant | 15 | 2.8 |
| Nurse Consultant | 7 | 1.3 |
| *Missing* | 16 |  |
| **Have you cared for any patients in the community who have died with confirmed (by test) COVID-19? (n=557)** |  |  |
| Yes | 296 | 53.1 |
| No | 261 | 46.9 |
| *Missing* | 2 |  |
| **Have you cared for any patients in the community who have died with suspected COVID-19 (untested but with clinical symptoms)? (n=554)** |  |  |
| Yes | 371 | 67.0 |
| No | 183 | 33.0 |
| *Missing* | 5 |  |
| **Have you been involved in providing end-of-life care at home for patients who do not have COVID-19 or suspected COVID-19 through the pandemic? (n=554)** |  |  |
| A lot more than usual | 172 | 31.1 |
| A little bit more than usual | 150 | 27.1 |
| About the same as usual | 211 | 38.1 |
| A little bit less than usual | 13 | 2.3 |
| A lot less than usual | 8 | 1.4 |
| *Missing* | 5 |  |
| **Have your working hours changed in order to deliver end of life care during COVID-19? (n=554)** |  |  |
| Yes | 189 | 34.1 |
| No | 366 | 65.9 |
| *Missing* | 4 |  |

### Theme 1: Increased need, complexity and unpredictability in community end-of-life care during COVID-19

Respondents reported increased involvement in the provision of end-of-life care in the community as a consequence of the pandemic, with over half providing end-of-life care ‘a lot more’ or ‘a bit more than usual’ (322, 58.2%) to patients who were not known or suspected to have COVID-19. Over half of respondents (296, 53.1%) reported caring for patients who had died with ‘confirmed’ COVID-19, and over two-thirds (371, 67.1%) reported caring for those who had died with ‘suspected’ COVID-19 (Table 1).

Working hours changed in response to the need, with these changes often arranged informally, and frequently unpaid.

> *‘‘Informal changes to work patterns, working well over shift times due to symptom management and workload also factoring in staff shortages due to COVID-19 and shielding” (Respondent 214, Community Staff Nurse, Northern Ireland)’*

An important factor was patients choosing to remain at home to receive end-of-life care rather than being admitted to hospital. This presented workload challenges, but was recognised as a long-term aspiration for patient care, achieved because of the pandemic:

> *“I feel more patients stayed at home for non-COVID related end-of-life care. Which was good. Think the staff that were at the front line went above and beyond to keep patients at home. Patients and families did not want admission as then they could not see family etc. and then die without family there. Staying at home was seen as best option for most patients and families, even if it was tiring”. (Respondent 55, District Nurse, England)*

Nurses highlighted an associated increase in the management of patients with more complex healthcare needs, not related to COVID-19:

> *“We have had more complex patients being managed at home which has been a challenge, whereas if COVID-19 and visiting wasn’t an issue they may have been hospice inpatients or even admitted to an acute hospital bed”. (Respondent 299, Sessional GP, England)*

Increased patient care needs resulting from the pandemic included an increase in frailty amongst those living with advanced illness who were shielding at home:

> *“Patients within the community have shielded well but we are seeing declining health conditions due to shielding … more frailty has been identified during COVID-19 as patients have lost their daily routines & independence. Lack of exercise and carrying out their normal activities of daily living has resulted in more frailty”. (Respondent 483, District Nurse Team Leader, England)*

The care of patients with COVID-19, as a new condition (or suspected COVID-19 when testing was not available), was associated with a high level of unpredictability and clinical uncertainty, and a variety of presentations of dying. Only four respondents reported patients dying with distressing symptoms due to COVID-19, such as agitation, breathlessness and abdominal pain. There were over 100 accounts of experiences of providing care for a large number of patients who deteriorated and died very rapidly, particularly frail elderly patients, including those in care homes:

> *“Before testing of clients I found that community and care home clients would be walking and healthy, then suddenly develop a temperature over 38 degrees [centigrade] and take to their bed, they would be lethargic and confused, not eat or drink and within approx 72 hours or so would have died.’’ (Respondent 142, Community Staff Nurse, England)*

### Theme 2: The roles of the primary care multi-disciplinary team to meet increased demand in the context of COVID-19

There were rapid changes in the roles of members of the community multi-disciplinary team (MDT) to meet the increased need for end-of-life care, with respondents reporting that they were undertaking more advance care planning (48.3%), anticipatory prescribing of medication (42.4%), symptom management (50.0%), bereavement support (43.1%) and death verification (41.2%) (Figure 1). Additionally, 339 (60.9%) reported that they were providing support to family and carers ‘a lot’ or ‘a bit more than usual’. There was variation in team-working activities with specialist palliative care teams, with 218 (39.5%) respondents reporting increased collaborative activities whereas 73 (13.2%) reported less.

**FIGURE 1.**
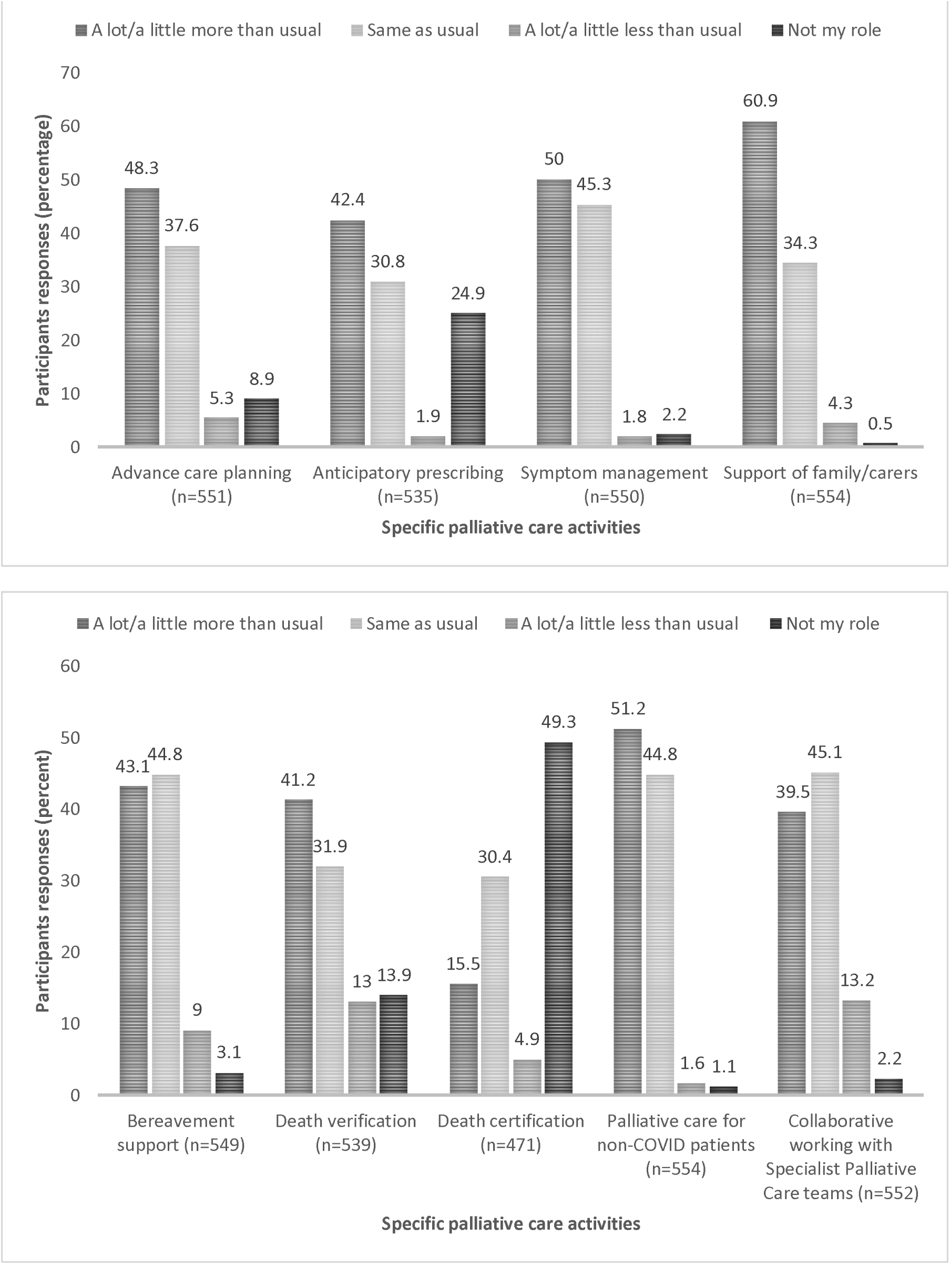
CHANGES IN PROFESSIONAL ROLE IN END-OF-LIFE CARE (N=559)

Comparing GPs and community nurses’ responses showed differences in their professional roles for palliative care activities (Table 2). GPs undertook ‘more’ or ‘a lot more’ advance care planning compared with the community nurses (p<0.0001), while community nurses reported providing ‘more’ or ‘a lot more’ symptom management (p<0.0001), bereavement support (p<0.0001), death verification (p<0.0001), palliative care for those without COVID-19 (p<0.0001) and collaborative working with specialist palliative care teams (p<0.0001). Although both groups indicated they had given more support for family and carers, a significantly larger proportion of community nurses answered ‘more’ or ‘a lot more’ compared with GPs (66.0% versus 48.1%, p=0.01).

**Table 2.**
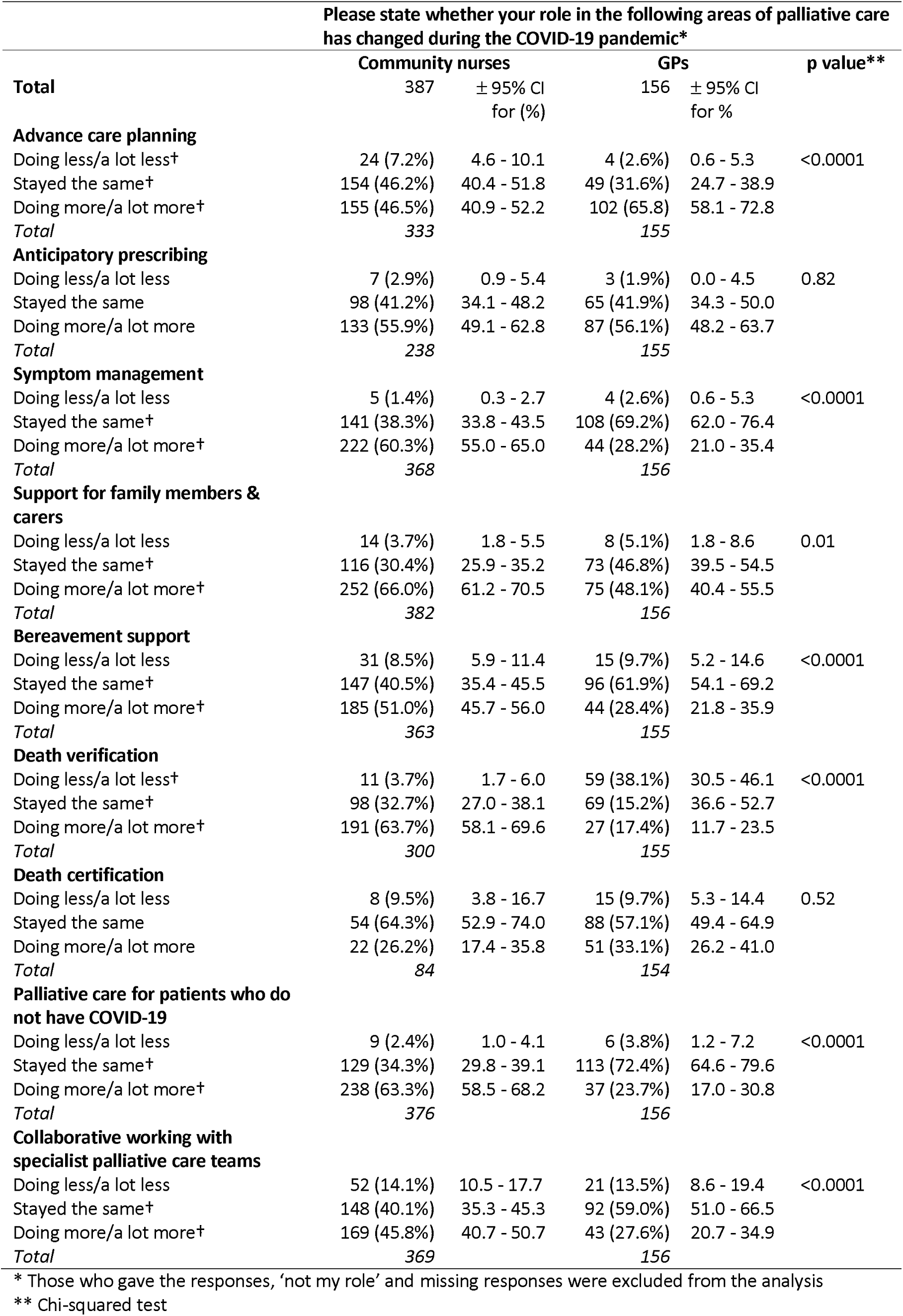
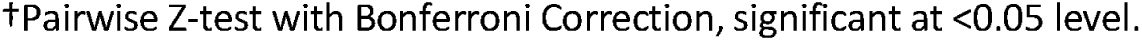
Comparison between community nurses and doctors’ responses for their professional role in palliative care activities.

| palliative care activities | Please state whether your role in the following areas of palliative care has changed during the COVID-19 pandemic* |  |  |  |  |
| --- | --- | --- | --- | --- | --- |
|  | Community nurses |  | GPs |  | p value** |
| Total | 387 | ± 95% CI<br>for (%) | 156 | ± 95% CI<br>for % |  |
| **Advance care planning** |  |  |  |  |  |
| Doing less/a lot less† | 24 (7.2%) | 4.6 - 10.1 | 4 (2.6%) | 0.6 - 5.3 | <0.0001 |
| Stayed the same† | 154 (46.2%) | 40.4 - 51.8 | 49 (31.6%) | 24.7 - 38.9 |  |
| Doing more/a lot more† | 155 (46.5%) | 40.9 - 52.2 | 102 (65.8) | 58.1 - 72.8 |  |
| Total | 333 |  | 155 |  |  |
| **Anticipatory prescribing** |  |  |  |  |  |
| Doing less/a lot less | 7 (2.9%) | 0.9 - 5.4 | 3 (1.9%) | 0.0 - 4.5 | 0.82 |
| Stayed the same | 98 (41.2%) | 34.1 - 48.2 | 65 (41.9%) | 34.3 - 50.0 |  |
| Doing more/a lot more | 133 (55.9%) | 49.1 - 62.8 | 87 (56.1%) | 48.2 - 63.7 |  |
| Total | 238 |  | 155 |  |  |
| **Symptom management** |  |  |  |  |  |
| Doing less/a lot less | 5 (1.4%) | 0.3 - 2.7 | 4 (2.6%) | 0.6 - 5.3 | <0.0001 |
| Stayed the same† | 141 (38.3%) | 33.8 - 43.5 | 108 (69.2%) | 62.0 - 76.4 |  |
| Doing more/a lot more† | 222 (60.3%) | 55.0 - 65.0 | 44 (28.2%) | 21.0 - 35.4 |  |
| Total | 368 |  | 156 |  |  |
| **Support for family members & carers** |  |  |  |  |  |
| Doing less/a lot less | 14 (3.7%) | 1.8 - 5.5 | 8 (5.1%) | 1.8 - 8.6 | 0.01 |
| Stayed the same† | 116 (30.4%) | 25.9 - 35.2 | 73 (46.8%) | 39.5 - 54.5 |  |
| Doing more/a lot more† | 252 (66.0%) | 61.2 - 70.5 | 75 (48.1%) | 40.4 - 55.5 |  |
| Total | 382 |  | 156 |  |  |
| **Bereavement support** |  |  |  |  |  |
| Doing less/a lot less | 31 (8.5%) | 5.9 - 11.4 | 15 (9.7%) | 5.2 - 14.6 | <0.0001 |
| Stayed the same† | 147 (40.5%) | 35.4 - 45.5 | 96 (61.9%) | 54.1 - 69.2 |  |
| Doing more/a lot more† | 185 (51.0%) | 45.7 - 56.0 | 44 (28.4%) | 21.8 - 35.9 |  |
| Total | 363 |  | 155 |  |  |
| **Death verification** |  |  |  |  |  |
| Doing less/a lot less† | 11 (3.7%) | 1.7 - 6.0 | 59 (38.1%) | 30.5 - 46.1 | <0.0001 |
| Stayed the same† | 98 (32.7%) | 27.0 - 38.1 | 69 (15.2%) | 36.6 - 52.7 |  |
| Doing more/a lot more† | 191 (63.7%) | 58.1 - 69.6 | 27 (17.4%) | 11.7 - 23.5 |  |
| Total | 300 |  | 155 |  |  |
| **Death certification** |  |  |  |  |  |
| Doing less/a lot less | 8 (9.5%) | 3.8 - 16.7 | 15 (9.7%) | 5.3 - 14.4 | 0.52 |
| Stayed the same | 54 (64.3%) | 52.9 - 74.0 | 88 (57.1%) | 49.4 - 64.9 |  |
| Doing more/a lot more | 22 (26.2%) | 17.4 - 35.8 | 51 (33.1%) | 26.2 - 41.0 |  |
| Total | 84 |  | 154 |  |  |
| **Palliative care for patients who do not have COVID-19** |  |  |  |  |  |
| Doing less/a lot less | 9 (2.4%) | 1.0 - 4.1 | 6 (3.8%) | 1.2 - 7.2 | <0.0001 |
| Stayed the same† | 129 (34.3%) | 29.8 - 39.1 | 113 (72.4%) | 64.6 - 79.6 |  |
| Doing more/a lot more† | 238 (63.3%) | 58.5 - 68.2 | 37 (23.7%) | 17.0 - 30.8 |  |
| Total | 376 |  | 156 |  |  |
| **Collaborative working with specialist palliative care teams** |  |  |  |  |  |
| Doing less/a lot less | 52 (14.1%) | 10.5 - 17.7 | 21 (13.5%) | 8.6 - 19.4 | <0.0001 |
| Stayed the same† | 148 (40.1%) | 35.3 - 45.3 | 92 (59.0%) | 51.0 - 66.5 |  |
| Doing more/a lot more† | 169 (45.8%) | 40.7 - 50.7 | 43 (27.6%) | 20.7 - 34.9 |  |
| Total | 369 |  | 156 |  |  |

\* Those who gave the responses, 'not my role' and missing responses were excluded from the analysis
\*\* Chi-squared test
†Pairwise Z-test with Bonferroni Correction, significant at <0.05 level.

**Table 3.** Comparison between community nurses and GPs responses for the type of consultation conducted.

| Please state whether your role in the following areas of palliative care has changed during the COVID-19 pandemic* |  |  |  |  |  |
| --- | --- | --- | --- | --- | --- |
|  | Community nurses | ± 95% CI for (%) | GPs | ± 95% CI for (%) | p value** |
| **Total** |  |  |  |  |  |
| **Home visits** | 387 |  | 156 |  |  |
| Doing less/a lot less† | 97 (44.5%) | 38.0 - 50.5 | 35 (28.9%) | 21.0 - 36.9 | <0.0001 |
| Stayed the same† | 36 (16.5%) | 11.5 - 21.5 | 66 (54.5%) | 45.1 - 63.2 |  |
| Doing more/a lot more† | 85 (39.0%) | 32.6 - 45.9 | 20 (16.5%) | 10.2 - 23.5 |  |
| *Total* | *218* |  | *121* |  |  |
| **Telephone reviews** |  |  |  |  |  |
| Doing less/a lot less† | 93 (37.2%) | 31.1 - 42.9 | 72 (50.7%) | 42.1 - 59.0 | p=0.08 |
| Stayed the same† | 7 (2.8%) | 0.8 - 5.2 | 0 (0.0%) | - |  |
| Doing more/a lot more† | 150 (60.0%) | 54.2 - 66.0 | 70 (49.3%) | 41.0 - 57.9 |  |
| *Total* | *250* |  | *142* |  |  |
| **Virtual consultations** |  |  |  |  |  |
| Doing less/a lot less† | 34 (13.0%) | 9.0 - 17.6 | 42 (34.1%) | 25.8 - 43.0 | <0.0001 |
| Stayed the same | 9 (3.4%) | 1.5 - 5.6 | 2 (1.6%) | 0.0 - 4.3 |  |
| Doing more/a lot more† | 218 (83.5%) | 78.8 - 88.1 | 79 (64.2%) | 55.2 - 72.6 |  |
| *Total* | *261* |  | *123* |  |  |
| **Face-to-face visits** |  |  |  |  |  |
| Doing less/a lot less† | 38 (13.9%) | 10.0 - 18.2 | 53 (39.8%) | 31.8 - 48.0 | <0.0001 |
| Stayed the same† | 25 (9.1%) | 5.8 - 12.8 | 79 (59.4%) | 50.8 - 67.6 |  |
| Doing more/a lot more† | 211 (77.0%) | 71.7 - 81.9 | 1 (0.8%) | 0.0 - 2.4 |  |
| *Total* | *274* |  | *133* |  |  |

\*those who gave the responses, 'not my role' and missing responses were excluded from the analysis
\*\* Chi-squared test
†Pairwise Z-test with Bonferroni Correction, significant at <0.05 level

Both GPs and community nurses reported an increase in consultations (telephone and virtual consultations and home visits) related to end-of-life care, but there were changes in the types of consultations undertaken (Figure 2). The greatest increase was in virtual consultations, with 308 (64.6%) undertaking ‘more’ or ‘a lot more’ of this type of interaction. There was a contrast between the two groups, with over three-quarters (77.0%) of community nurses reporting ‘more’ or ‘a lot more’ face-to-face visits, whereas almost 40% of GPs reported they were doing ‘less’ or ‘a lot less’ (p<0.0001).

**FIGURE 2.**
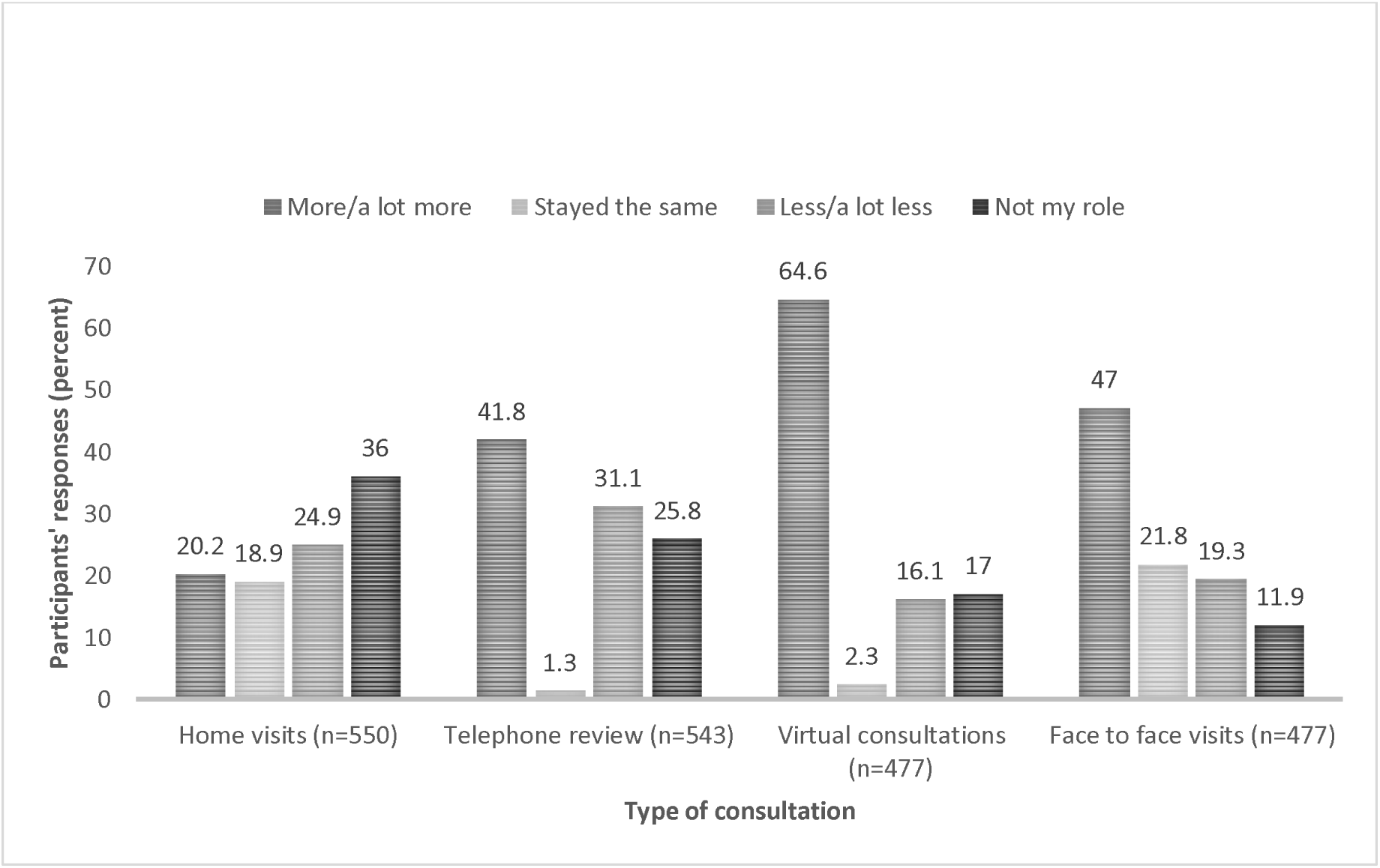
CHANGES IN TYPE OF CONSULTATION CONDUCTED.

Both GPs and community nurses, described concerns about remote consultations in end-of-life care. There were descriptions of a loss of “*professional intimacy*” *(respondent 37, GP partner, England),* and frustrations amongst professionals about the limitations of telephone or virtual contacts with patients:

> *“I have also found it frustrating to be giving support by telephone when I would usually be visiting a patient to assess their needs. I am quite sure that some patients have deliberately played down their health issues because they felt that the nurses were too busy already and didn’t want to be a burden”. (Respondent 78, Community Staff Nurse, Scotland)*

It was felt that telephone and virtual consultations were a cause of confusion and distress for patients, and caused *“huge emotional trauma to families” (Participant 441, District Nurse, Northern Ireland)*

> *“The virtual contact was confusing for some patients and almost became something just to tick the box that you had ‘seen’ them in case they passed away”. (Respondent 498, GP Partner, England)*

A frequent response from community nursing staff was that while their visits to patients had increased, other services, both general practice and specialist palliative care services, had “*stepped back completely*” leaving them feeling abandoned, vulnerable and “*more disposable*” than other colleagues:

> *“GPs and other community services have backed away leaving us to deal with a lot of difficult situations and questions from families. I feel the end of life care given by GPs has been dangerous and neglectful and has not been seamless and high quality”. (Respondent 129, District Nurse, England)*

GP respondents shared this concern. Some GPs reported continuing to provide regular home visits for end-of-life care, but others described feeling under pressure to decrease their home visits due to infection control procedures, resulting in them not being able to “g*ive the support [to patients, families and nursing colleagues] I would normally wish to” (Respondent 475, Sessional GP, England).* The need for clinical leadership within the team providing end-of-life care in the community, while not unique to GPs, was compromised by service changes:

> *“we were encouraged as GPs not to visit care homes if possible, however our clinical leadership role within the care home setting was missed and when we got back in there it helped to get back to our usual multi-disciplinary team working … The idea that you can replace that sort of team with technology at a time of great stress was proved wrong”. Respondent 135, GP partner, Scotland.*

Views of support from specialist palliative care services were mixed, with some participants describing a positive opportunity to work more closely with these services in the community. Some experienced support through online communities of practice set up by their local hospice. However, others described difficulties communicating and co-ordinating care with specialist palliative care colleagues, describing them as “*distant*” and again reinforcing the reliance on community nursing staff:

> *“District Nurses have borne the bulk during COVID. They are, and continue to be, amazing. The ‘specialist’ palliative care nurses occasionally raise their heads, but are distant and remote from general practice which is a shame” (Respondent 67, Deputy District Nurse Team Leader, England)*

### Theme 3: Fears, repeated loss and undervaluation of contributions threatens resilience and wellbeing of community healthcare teams

Changes in the delivery of care, team dynamics and roles, and increased involvement in end-of-life care had consequences for the emotional wellbeing of respondents. Some described teams “*pulling together to get on with it”* and becoming “*a stronger, more bonded*” team. Others described their experiences as “*emotionally and physically draining*”. Witnessing high volumes of people dying, and managing anxiety around infection control, were significant factors:

> *“[Nursing] staff felt accused of bringing the virus into the home, there was some public shaming as well as public support, the issue of [personal protective equipment] clouded everything (was there enough, was it being used correctly, who was responsible?) … Deaths were more rapid than flu. Many staff got the virus but thankfully no one seriously … Staff have been left broken and there are symptoms of [post traumatic stress disorder], depression and anxiety” Respondent 148, GP partner, Scotland*

Respondents described the impact of staff shortages within the team as a result of colleagues becoming unwell with COVID-19, or having to self-isolate. Some had experienced the death of a colleague from COVID-19. Others expressed fears for their own safety particularly if they had health concerns of their own:

> *“I have been frightened coming to work for the last 5 months as I [have risk factors] so a prime candidate for COVID 19. But am proud to say I have not shirked my responsibility and have done my duty. It has been very difficult especially when rushing to someone poorly’s house and having to doff and don [*personal protective equipment*] before seeing the patient. (Respondent 149, District Nurse, England)*

Support from colleagues, including nursing team leaders was highly valued although formal psychological support was lacking. Many respondents described feeling that, as community and primary care staff, their contributions to patient care during COVID-19 did not receive the same attention as hospital care during COVID-19 due to a “*concerning focus on critical care capacity in hospital” (Respondent 1, Sessional GP, England)*. This had a negative impact and contributed to a perception that they were undervalued:

> *“….the general public seemed to think COVID-19 only existed in hospitals and not in people’s homes. Care homes were forgot about and they seemed to have little or no guidance in relation to infection prevention’’ (Respondent 200, District Nurse, England)*

## Discussion

### Summary of main findings

During the first wave of the COVID-19 pandemic, community nurses and GPs experienced a substantial increase in the need and complexity for palliative and end-of-life care. Specific palliative care activities increased, with community nurses taking greater responsibility in most areas of care including symptom control and the provision of support to family members. GPs reported an increase in advance care planning. Working hours changed to meet rising demands for care at home through a mainly ‘ad hoc’ approach.

Changes in the mode of service delivery, including increased virtual consultations, resulted in reports of disconnection within and between teams. Community nursing team members particularly described a sense of abandonment and perceived that other services, including general practice and specialist palliative care, had withdrawn. GPs reported feeling that the use of virtual consultations was limited in the end-of-life care context. A significant emotional toll was experienced due to the impact of providing care during COVID-19 with fears relating to uncertainty and loss of the usual mechanisms of interdisciplinary and collegial support.

### Strengths and limitations

This is the first UK-wide survey undertaken to understand the impact of the COVID-19 pandemic on healthcare services involved in the provision of community-based end-of-life care. It provides valuable insights into the role of primary healthcare and the findings are highly relevant to practice, service delivery models and policymaking as the pandemic progresses.

The minimum number of returns was achieved, but the response rate was low amongst GPs even though the survey was distributed through well-respected professional organisations. This may be due to the timing of the survey, when workload related to the pandemic was high and that there were a large number of other surveys taking place concurrently. The findings are likely to reflect the views of primary care professionals with an active role or interest in palliative care and may not be representative of the wider population of GPs and community nurses.

### Comparison with existing literature

There is a lack of previous research to inform practice, service delivery or policy (9, 16) despite the importance of the provision of high quality end-of-life care in a pandemic (17, 18). Respondents in this study perceive that their role in the pandemic response has received less focus than the response of hospital care (19, 20). Primary healthcare teams have a pivotal role in end-of-life care, and feeling undervalued may have contributed to the significant emotional distress experienced during COVID-19.

The increased need for palliative and end-of-life care in the community is in keeping with research from previous pandemics and is borne out in COVID-19 population data (1, 21). The increase in deaths in the community has placed extra time and resource pressures on both GPs and community nurses, often met through the efforts of highly committed individual healthcare professionals (19). Much of the increase in care was for patients dying of conditions other than COVID-19 as a result of patient choice to stay at home, particularly with visiting restrictions in hospitals (22). The resulting increase in delivery of end-of-life care and management of complexity in the community contrasts with fears at the start of the pandemic that end-of-life care would be required for a large number of people dying with COVID-19, with such significant pressure on hospital beds, including critical care, that admission would have to be avoided.

Community nurses have previously described end-of-life care as one of the most rewarding aspects of their job (23). This research provides insights into the new responsibilities rapidly assumed by community nurses in almost every aspect of end-of-life care during COVID-19. The context for the delivery of end-of-life care in the community changed drastically with national lockdowns, self-isolation and shielding resulting in patients at home alone in the community, unable to access their usual social support. Community nurses represent one of the few professional groups that people at the end-of-life had face-to-face contact with during the pandemic. The level of skill, compassion and resilience required to undertake this task must be recognised and valued (24).

Previous research has exposed tensions in relationships both within primary healthcare teams and between these teams and specialist palliative care colleagues (25). Increased use of virtual consultations by GPs and specialist palliative care teams during COVID-19 appeared to exacerbate such conflicts, with community nurses describing a feeling of abandonment and isolation. The interpretation of policies recommending that home visits by GPs should be limited as far as possible (26) may have contributed to this situation. GP respondents in this survey and elsewhere have specifically highlighted the importance of face-to-face home visits as an ongoing priority for care of vulnerable patients in the community, including those who are dying (27, 28), and described a sense of moral distress related to the change in their role associated with increased use of remote consultation. The role of GPs working alongside community nurses in the provision of end-of-life care not only involves specific clinical tasks, but also reviewing, affirming and supporting care decisions within the multi-disciplinary team. This is particularly relevant in the context of COVID-19 as a new condition, where clinicians must take responsibility for complex and nuanced clinical decision-making, and collectively manage a great deal of clinical risk and uncertainty (29).

Previous research has demonstrated the profound emotional impact that providing care for the dying during a pandemic can have on staff, and recommended access to training, support and debriefing as part of a pandemic response (30). This remains relevant and it is important to recognise.

### Implications for practice, policy and research

This survey provides valuable insights into experiences of professionals during the first wave of the pandemic, highlighting the many changes that occurred and the impact on both services and individuals. Future research should include more in-depth investigation into specific aspects of end-of-life care such as advance care planning and end-of-life care in care homes. There is much more to learn about the impact and role of remote consultations in end-of-life care if this practice is to continue. Further analysis of qualitative responses yielded in this survey is planned. Research to understand the perspective of patients and families currently experiencing palliative and end-of-life care would be valuable. This further research is required in the context of a constantly evolving pandemic situation and changing knowledge of the management of COVID-19 which has a direct impact on care decisions such as whether or not a patient would benefit from admission to hospital.

There is an immediate need for policy makers and commissioners to recognise the sustained increased need for end-of-life care in the community and the critical role of primary healthcare services in the delivery of this care as the pandemic progresses. Policy guidance and service models must place focus on and support the multi-disciplinary team relationships that are necessary to deliver this care most effectively, rather than the roles of specific services as this has the potential to fragment teams. Ensuring support for individuals involved in the provision of this care, through team relationships, training opportunities and debrief also requires attention. The findings of the survey suggest a disconnect between teams involved in end-of-life care in the community and a need to rebuild trusted relationships through truly integrated approaches between GPs, community nurses and specialist palliative care services.

## Conclusion

This study has identified the contrasting and potentially conflicting roles that emerged between GPs and community nurses in their responses to the increased demand and complexity of palliative and end-of-life care in the community in the early months of the COVID-19 pandemic. The significant emotional impact, especially for community nurses, needs to be addressed alongside promoting effective, collaborative and mutually supportive team working that can recognise and quickly adapt to changing patient needs.

## Data Availability

Data are available upon reasonable request to the corresponding author.

## Ethical approval

Ethical approval was granted by the University of Sheffield Research Ethics Committee (UREC) Ref No: 035508. Approved 28.7.20

## Authorship Statement

SM, CM, JD, KB and SB conceptualised the study. CG and DK led the PPI work. All authors contributed to the survey dissemination and data collection. SM, CM and PO led the data analysis. SM and CM drafted the article. CG, HC, DK, JD, JB and SB reviewed the article critically for clarity and intellectual content and provided edits and revisions. All authors have approved this version for submission.

## Acknowledgements

The authors would like to thank all of the GPs and community nurses who took part in this survey study.

